# Protocol for an EHR-embedded pragmatic randomized control trial of Ambient AI to Reduce Nursing Staff Documentation Time

**DOI:** 10.64898/2026.07.09.26357653

**Authors:** Ann Wieben, Jann Pfaff, Mary Ryan Baumann, Felice Resnik, Sarah Brzozowski, Chelsey Langer, Kristen Stine, Colin Gillis, Anne Gravel Sullivan, Caitlin Voegele, Leigh A Mrotek, Majid Afshar, Elizabeth S. Burnside, Jennifer Hankwitz, Stacy Rasmussen, Rudy Jackson, Becky Kohler

## Abstract

**Background:** Documentation burden significantly impacts nursing workload and well-being, with nurses spending an estimated 20-40% of their time on documentation. Ambient AI technologies offer potential to reduce documentation time by mapping real-time nurse-patient conversations to structured EHR data entries with human-in-the-loop verification.

**Methods:** This protocol describes a pragmatic, EHR-embedded randomized controlled trial evaluating the effectiveness of an Ambient AI tool in reducing nursing documentation time across three inpatient medical/surgical units. The study employs a closed-cohort, stepped-wedge, unit-randomized design, integrating the intervention into routine clinical workflows. The primary outcome is documentation time per shift hour, derived from EHR audit logs. Secondary outcomes include documentation burden, professional well-being, and perceived usability.

**Results:** The trial is being implemented within a shared governance model that integrates executive oversight, operational feasibility, and research rigor. Multidisciplinary workgroups coordinate technical integration, user experience, and analytics, ensuring alignment between operational priorities and pragmatic trial objectives. Early implementation has highlighted the importance of adapting training and analytic strategies to address differential intervention exposure, as well as the need for rapid operational responses to late-emerging technical issues.

**Discussion:** This protocol demonstrates the feasibility of embedding a randomized pragmatic trial within a health system-led operational deployment of Ambient AI for inpatient nursing documentation. The approach highlights the necessity of adapting existing outpatient provider-focused AI implementation strategies for inpatient nursing, emphasizing the unique nature of different nursing care environments. Recruitment challenges and the integration of research with operational workflows are discussed as key considerations for future pragmatic AI trials in nursing.

**Funding:** The protocol development was funded by University of Wisconsin Hospitals and Clinics (UW Health) and UW Institute for Clinical and Translational Research Learning Health System

**ClinicalTrials.gov Identifier:** NCT07456241V4: 2026-05-27 https://clinicaltrials.gov/study/NCT07456241

## Introduction

### Background and Rationale

Nursing documentation is a time-intensive activity that occupies up to 40% of a nursing shift [1]. Documentation in flowsheets and other structured data fields contributes significantly to this burden; prior studies found nurses complete 631-875 manual data entries [2], consuming well over two hours of worktime per shift [3]. Documentation burden in nursing can cause stress and increase an already heavy workload, especially when documentation systems fail to support care delivery. Nurses consistently perceive their documentation workload as high [4], and this burden has been associated with emotional exhaustion and depersonalization [5]. Electronic Health Record (EHR)-stress is also prevalent, with nurses reporting moderate to high levels [6]. EHR-stress has been characterized as stress experienced by clinicians due to EHR design and use factors that interfere with patient care or create inefficient systems [7]. Dissatisfaction with documentation system usability is well established, generating interest in alternative documentation strategies to alleviate nursing documentation burden and the associated negative effect on nurse well-being [8–10]. These consequences have implications not only for individual well–being, but also health-system-level staffing stability and workforce sustainability.

Ambient Listening technology, or Ambient AI, may reduce documentation burden by leveraging automated speech recognition (ASR) and large language models (LLMs) to transform real-time clinical conversations into drafted EHR documentation records [11]. Ambient AI has been used more widely for healthcare practitioner workflows, showing promise in reducing time spent in documentation, improving healthcare practitioner well-being and communication with patients [12]. A recent literature review identified only ten studies that examined the impact of speech recognition tools on nursing documentation measures (e.g., documentation quality, documentation time, acceptance), however, few of the included studies employed a technology that mapped speech to discrete documentation fields [13]. Rigorous empirical evaluation is still needed to formally assess this technology’s usability, effectiveness, and impact on nursing workflows, particularly in busy acute care hospital settings. Objective, EHR–derived metrics can be particularly important for evaluating documentation technologies in inpatient nursing contexts, because they enable continuous, unbiased measurement of documentation time and work patterns. EHR-derived metrics are well suited to informing operational decision–making within a learning health system (LHS) framework [14]. The purpose of this paper is to outline the collaborative development of a pragmatic randomized controlled trial and provide a playbook for the implementation and evaluation of Artificial Intelligence (AI) facilitated nursing documentation technology.

## Methods

### Trial design

This study uses a pragmatic randomized controlled trial with a stepped–wedge design to evaluate Ambient AI technology under routine clinical care. Nursing staff (registered nurses and nursing assistants) across three inpatient units at an academic medical center are randomized to staggered intervention start times as part of a health system-initiated operational Ambient AI deployment. Embedding the intervention within the EHR supports integration into existing workflows and enables evaluation of the intervention’s effectiveness over time while aligning with health system implementation needs. The impact of the intervention will be evaluated using administrative nursing documentation metrics across all study units. Nursing staff who provide informed consent will additionally contribute individual–level documentation metrics and participate in surveys, with optional interviews, focused on perceived EMR–related stress and system usability.

### Study Setting

This study will be conducted on three Medical/Surgical inpatient units at a Midwest academic medical center. Each unit comprises 27-28 beds and serves a mixed medical–surgical patient population, with standard staffing ratios of 4-6 patients per registered nurse and 8-10 patients per nursing assistant. Currently, nursing staff manually document assessment findings, measurements, and care provided in the EHR via discrete data fields structured in flowsheets with some textual notes/comments. Across the three medical/surgical units, most documentation is completed on three distinct flowsheets. The number of rows per flowsheet varies but ranges from a minimum of 45 to a maximum of 87 (with the ability to add rows not visible by default). Therefore, nursing staff typically tap/click/scroll from top to bottom.

### AI Intervention and Clinical Practice Integration

The AI intervention consists of the Abridge™ Ambient AI tool, a multi–component AI solution that incorporates ASR and LLMs. The system uses a smartphone microphone to capture real–time nurse-patient conversations and interactions, which are transcribed and processed to generate structured documentation (Figure 1). AI–generated outputs consist of discrete EHR flowsheet values that are presented to nursing staff for review. All AI-drafted documentation requires nursing staff confirmation before finalized entry into the permanent medical record, and nursing staff may edit or delete any AI–generated content deemed inaccurate or inappropriate. Documentation that is not confirmed by nursing staff is not filed, ensuring that final clinical documentation reflects nursing staff judgment and accountability.

**Figure 1.** Representative screenshots illustrating the Ambient AI tool interface and related EHR flowsheet documentation.

The Abridge Ambient AI tool was embedded within Epic^®^ EHR via the Rover mobile application installed on handheld devices that were already in place for healthcare operations, enabling secure and workflow–aligned use at the point of care. The Ambient AI tool was implemented under an established software–as–a–service vendor contract.

The Ambient AI tool is designed to generate structured flowsheet entries only when supported by the underlying nurse–patient conversation, with multiple safeguards in place to help prevent unsupported draft documentation (Figure 1). Previous validation was conducted by the vendor using annotated datasets reviewed internally by Abridge registered nurses, and model outputs are further tested in partnership with health systems prior to and during deployment.

### Outcomes and Endpoints

Primary and secondary outcomes are summarized in Table 1. The primary objective is to evaluate the effect of Ambient AI on nursing documentation time in flowsheets. The primary outcome is nursing documentation time, measured as minutes spent in EHR flowsheets per shift hour. This outcome is clinically relevant given the well–established association between documentation burden, workflow inefficiency, after–hours charting, and clinician burnout. A secondary objective is to assess the impact of Ambient AI on nursing documentation burden. Secondary outcomes capture complementary dimensions of documentation burden and user experience. These include documentation time per patient per shift, number of clicks or taps in flowsheets, and documentation time outside assigned shift hours. These primary and secondary outcomes are derived from the EHR audit log data.

**Table 1.** Summary of primary and secondary outcomes with associated metrics.

| Outcomes | Metrics |
| --- | --- |
| **Primary** |  |
| Nursing staff documentation time | Active Time Spent in Flowsheets per shift hour |
| **Secondary** |  |
| Nursing staff documentation burden | • Active time spent in flowsheets per patient per shift<br>• Clicks/taps in Flowsheets<br>• Time spent documenting after end time of assignment (i.e. overtime charting) |
| Nursing staff well-being | Professional Wellbeing (Mini-Z 3.0): Subscales of work conditions and EMR stress |
| Nursing staff perceived usability of documentation system | Staff perceived usability of the Ambient AI tool (SUS) |

Another secondary objective is to evaluate the association between ambient AI use and nursing staff professional well-being. For nursing staff who consent to participate in survey data collection, professional well–being and EHR–related stress are assessed using the Mini–Z (3.0) survey [15] with differences in total and subscale scores evaluated between conditions. An additional secondary objective is to assess nursing staff perceived usability of the Ambient AI tool. Perceived usability of the Ambient AI tool is measured using the System Usability Scale (SUS) [16], summarized as mean scores and compared against established benchmarks for satisfactory usability. Together, these outcomes will provide clinically meaningful evidence of whether the Ambient AI intervention reduces documentation burden while supporting usability and staff professional well–being, which are key criteria for the safe and effective implementation of AI in routine clinical practice.

### Eligibility Criteria

Eligible participants for individual–level analyses are registered nurses and nursing assistants aged ≥18 years who are English–speaking, trained in Ambient AI use, and not planning extended leave (≥6 weeks) during the calendar year.

### Recruitment

Potentially eligible nursing staff were invited to participate in the survey and interview components of this study through direct email utilizing scripts approved by the health system and the local IRB. Flyers announcing that volunteers are needed for a study were posted in nursing staff common areas, such as break rooms.

### Consent

Nursing staff eligible for individual–level surveys and interviews will be invited via email and provided with a study information sheet. Participation is voluntary, with implied consent indicated by completion of the survey; participants may withdraw at any time without penalty. Patient consent for Ambient AI is incorporated into standard inpatient admission workflows and documented in the EHR, focusing on permission to record conversations rather than research participation, as nursing staff are the study subjects. Posted signage will inform patients of Ambient AI use, and functionality will be disabled if consent is declined. Capacity–based consent workflows will be used when applicable, including proxy consent for patients without capacity and parental consent for pediatric patients. Although patients are not study participants, de–identified patient data will be collected to describe the clinical context of care.

### Randomization

Nursing units will be randomized 1:1:1 to intervention implementation times. Each unit will transition from control to intervention at different time points according to the randomized schedule (Figure 2), allowing for a staggered implementation across the study period that aligns with the operational needs of onboarding and training nursing staff to the tool. Computer-generated randomization sequence will be used to assign nursing departments to different start times for the intervention (sequence). The randomization was performed by the study statistician who was not involved in the training of the participants. Following a 4–week standard–of–care period, units will transition to the intervention at 3–week intervals over 22 weeks. Given the nature of the intervention, blinding will not be feasible.

**Figure 2.**
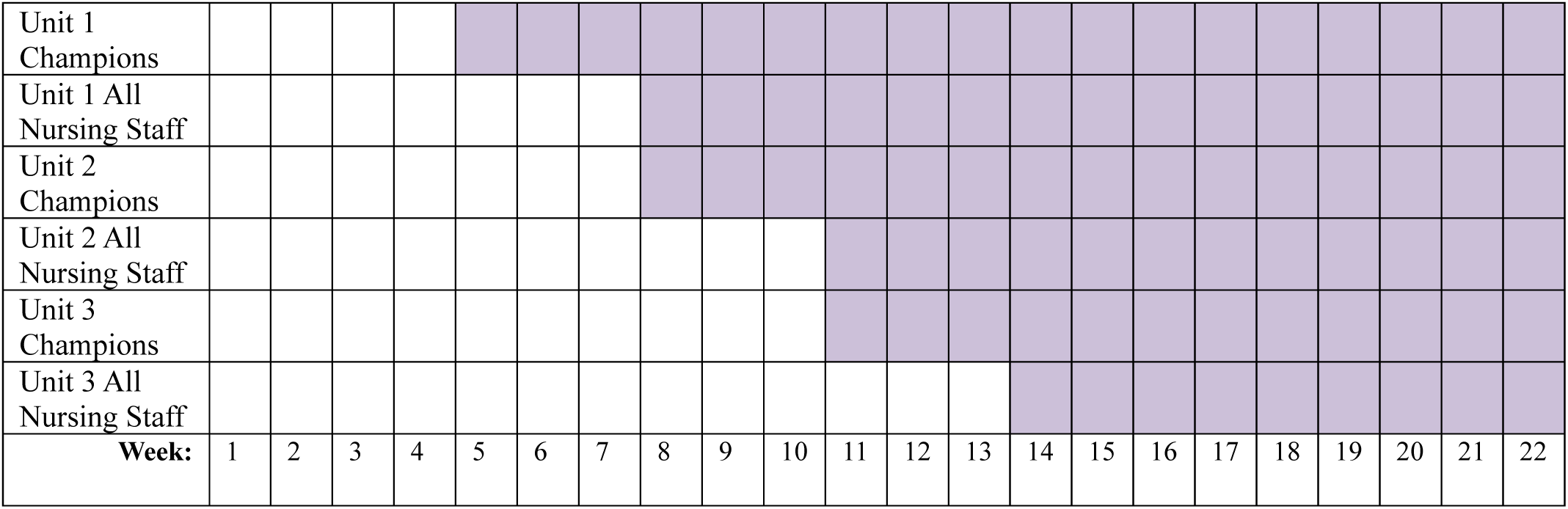
Randomized schedule showing staggered unit transitions from control to intervention, with nurse champion sub-waves, for evaluation of Ambient AI in nursing documentation

### Data Collection

Data collection will occur at baseline and at prespecified on–study and end–of–study time points using multiple data sources (Table 2). Quantitative data include EHR–derived documentation and usage metrics and survey–based measures of professional well–being and perceived usability of Ambient AI. Surveys will be administered approximately every three weeks. EHR audit logs will provide continuous data on documentation time, utilization, and interaction with AI–generated outputs and will be extracted regularly and securely transferred to the study biostatistician. Qualitative data from optional interviews and in–tool feedback will contextualize quantitative findings and capture nursing staff experiences. Baseline characteristics (e.g., demographics, clinical role, work hours) will also be collected via a baseline survey. EHR and vendor–generated data will be obtained through health system enterprise systems, survey data will be managed in REDCap (Research Electronic Data Capture)[17], and vendor–provided adoption metrics will be used without retention of audio recordings.

**Table 2.**
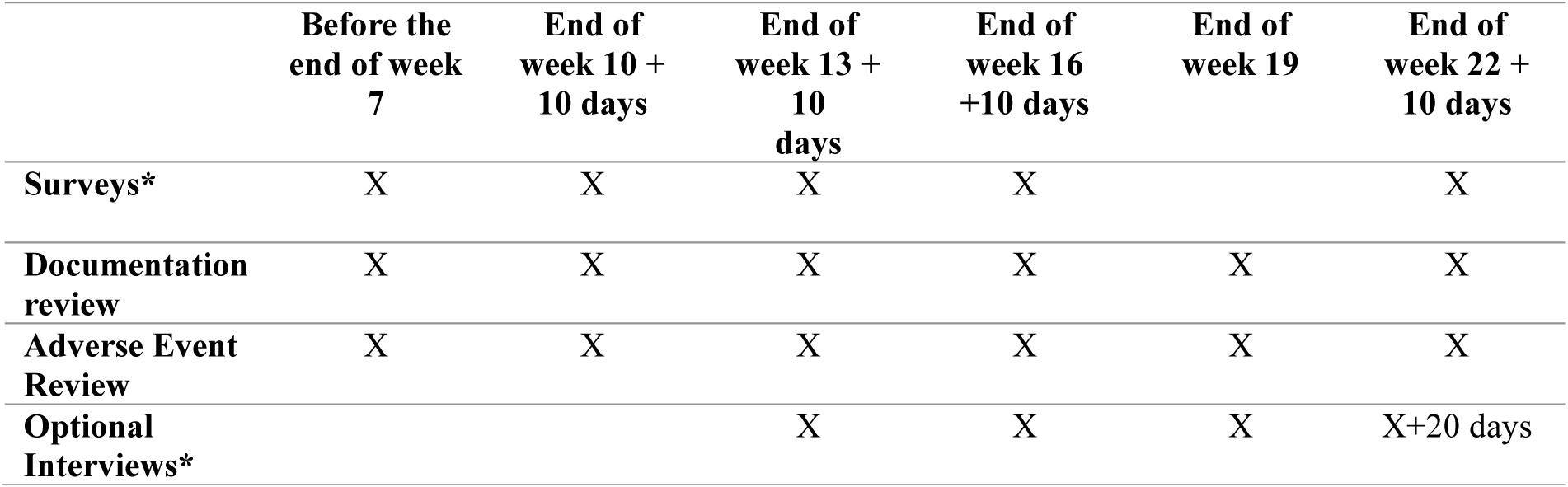
Timepoints for study procedures. Procedures requiring nursing staff consent are marked with *.

|  | **Before the**<br>**end of week**<br>**7** | **End of**<br>**week 10 +**<br>**10 days** | **End of**<br>**week 13 +**<br>**10**<br>**days** | **End of**<br>**week 16**<br>**+10 days** | **End of**<br>**week 19** | **End of**<br>**week 22 +**<br>**10 days** |
| --- | --- | --- | --- | --- | --- | --- |
| **Surveys\*** | X | X | X | X |  | X |
| **Documentation**<br>**review** | X | X | X | X | X | X |
| **Adverse Event**<br>**Review** | X | X | X | X | X | X |
| **Optional**<br>**Interviews\*** |  |  | X | X | X | X+20 days |

### Sample Size

Historical data for the primary outcome indicate a standard deviation of 3 minutes for flowsheet time. Sample size was initially estimated assuming four waves with an average of 52 nursing staff per wave to accommodate the three nursing units and a fourth non-randomized cluster of nurse champions. Assuming a within-period unit-level intracluster correlation (ICC) of 0.05, a unit autocorrelation of 0.8 (between-period ICC of 0.04), a within-nursing staff member ICC of 0.8, a two-period decay correlation structure, and a two-sided 5% type I error, the study will have 84% power to detect a 24-second reduction in documentation (time spent in EHR flowsheets per shift hour). This effect size was selected as clinically meaningful because even modest time savings per documentation episode may accumulate substantially across repeated patient-days and nursing workflows. If the standard deviation is as large as 4 minutes, the study maintains 82% power to detect a 31.2-second reduction. Current administrative data indicate there are approximately 127 RNs and 72 nursing assistants who will potentially meet eligibility criteria for this study across the three units. It was later decided that it would be logistically more feasible for nurse champions to sequentially gain access to the intervention ahead of their respective nursing units, instead of together as a single champion unit; this creates six implementation sub-waves within three randomized parent waves. Thus, we may consider the above to be conservative sample size estimates.

### Statistical Analysis

We conduct this study using intent-to-treat (ITT) principles. We will assess the primary hypothesis at two levels of analyses, the individual nursing staff member and the nursing unit. For the individual-level analysis, the primary outcome (time spent in EHR flowsheets per shift hour) will be assessed via a linear mixed effects model with random effects for nursing unit and nursing staff member, as well as a random effect interaction between nursing unit and time period. Secular trend will be adjusted for via a continuous time since baseline linear fixed effect; nurse champion status will be adjusted for via a binary fixed effect. Small cluster adjustments will be used. A similar model will be used for the nursing unit-level analysis, where the primary outcome will be aggregated to the nursing unit-level average; only unit-level random effects will be included, but the model will still adjust for secular trends. The primary analysis will use a two-sided type I error rate of 0.05. Secondary outcomes will be assessed in a similar manner, where continuous outcomes will use an identity link and binary outcomes will use a logit link.

For the survey data, Likert endpoints (1-5 scale) will be dichotomized to “achieves 1-2 rating” or “achieves 4-5 rating”, depending on outcome sentiment (positive/negative). Models for secondary outcomes collected via REDCap survey will adjust for secular trend via categorial time period fixed effects. To control for false discovery for secondary outcomes, we will use the Benjamini Hochberg method with a total two-sided type I error of 0.1[18]. All instances of missing data will be documented. Sensitivity analyses will estimate effects under assumptions of Missing Completely at Random (MCAR), Missing at Random (MAR), and Missing Not at Random (MNAR).

### Data monitoring and Auditing

Monitoring will be performed by the clinical operations governance team, with study subject to suspension for unexpected or unacceptable risk of futility. To monitor for unexpected intercurrent events (e.g. software or EHR updates), the study biostatistician will perform a moving-window difference-in-differences analyses, as described in [19] using one-month observation windows updated bimonthly. Statistically significant deviations (Bonferroni-corrected P values ≤ 0.05) will trigger root cause analyses and review with operational leaders. In addition, nursing staff can rate and provide free–text feedback on each ambient recording output through an embedded interface, supporting real–time detection of issues and timely adjustments to the model or local technical build needs by vendor and informatics teams.

### Research Ethics Approval

This study protocol has been developed in accordance with the SPIRIT-AI guidelines to ensure transparency and rigor [20]. The protocol has been approved by the University of Wisconsin Institutional Review Board and the participating hospital’s Research Operations Committee.

### Confidentiality

Study participant confidentiality and privacy will be rigorously protected throughout the study. All investigators and study personnel will complete required training in human subjects’ protection, the Health Insurance Portability and Accountability Act (HIPAA), and Good Clinical Practice and will carry out the study accordingly.

### Data sharing and dissemination

This study will comply with applicable publication and data–sharing policies, including the NIH Public Access and Data Sharing policies. Peer–reviewed manuscripts will be submitted to PubMed Central upon acceptance, and the trial is registered on ClinicalTrials.gov (NCT07456241) with results reported as required. De–identified study data may be shared with qualified researchers following completion of the primary endpoint, subject to investigator and institutional approval. The study protocol, data dictionary, analysis code, and de–identified survey data will be publicly available at: https://git.doit.wisc.edu/smph-public/LearningHealthSystem/ambientlisteningnurseinpatient.

## Results

The trial is being implemented within a shared governance model designed to align executive oversight, operational feasibility, and research rigor (Figure 3). This was an enterprise-led initiative with a priority for operations workflows along with a Learning Health System approach to produce analytic rigor and actionable evidence. Governance consists of executive sponsors, a multidisciplinary oversight group, and three function–specific workgroups, supported by a project management dyad and integrated LHS team members. Executive sponsorship includes the Chief Nursing Informatics Officer (CNIO), an Information Services vice president, and the Chief Nursing Officer (CNO). This group provides strategic oversight, approves major design and implementation decisions, and maintains accountability for vendor relationships. Decisions made at the executive sponsor level included approval of trial design, scope of rollout, operational consent model, constraints on tool use (such as use for dictation outside of the patient room), and escalation of vendor–related issues.

**Figure 3.**
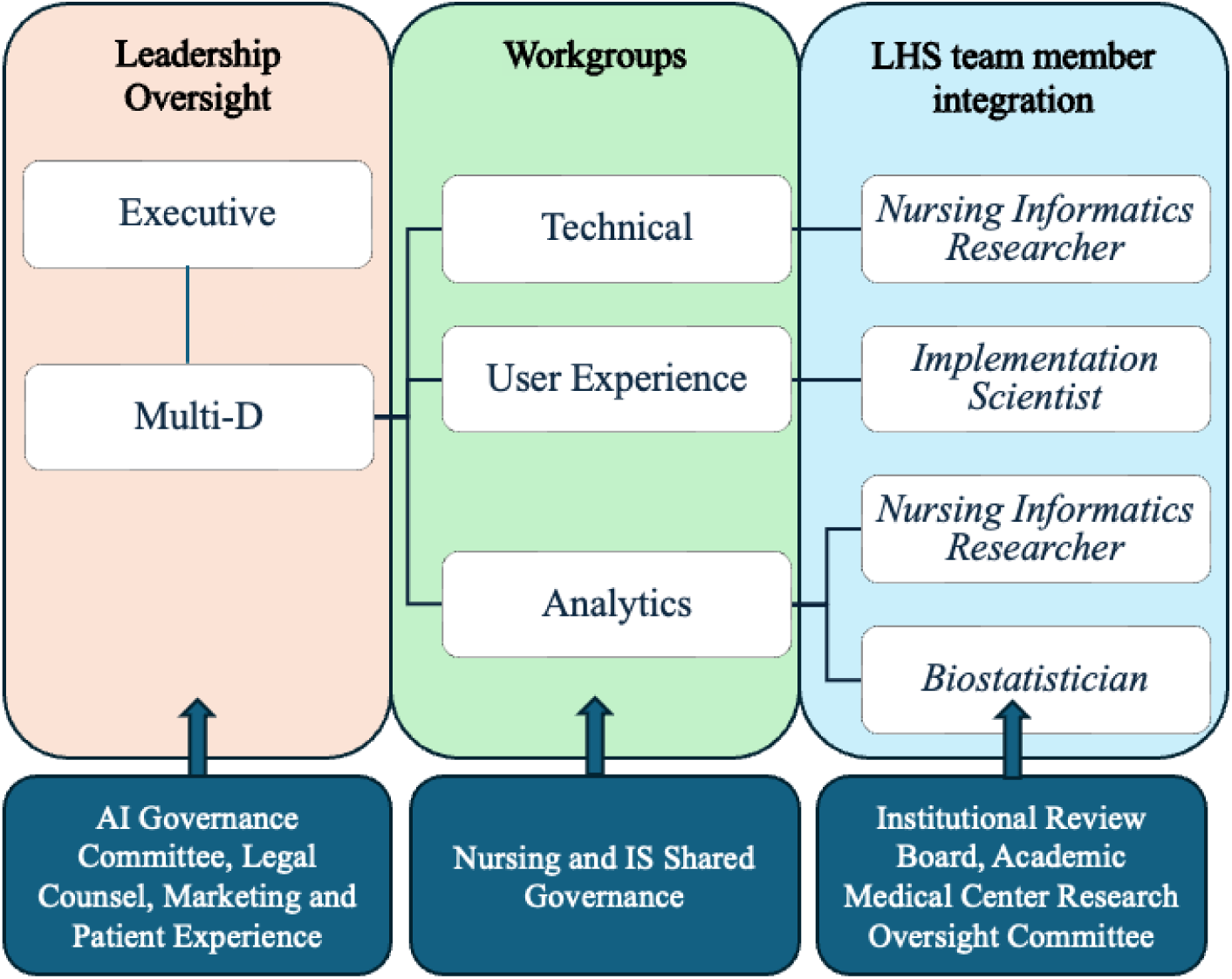
Governance structure with Pragmatic Trial Operations. Main project governance structure consisted of Leadership Oversight, Workgroups and LHS team member integration. Supporting organizational governance bodies providing approvals and/or consultation are also depicted.

A multidisciplinary oversight team is responsible for operational coordination and cross–functional decision–making throughout the study. This group includes executive leaders, workgroup leads, and legal counsel. Core responsibilities include coordination of project timelines, workflow review and change management strategy, marketing and patient engagement and consent approaches. Three workgroups were established to operationalize the intervention and evaluation: technical build, user experience/training, and analytics. The involvement of two nurse scientists (AW, JP) in all three workgroups strengthened conceptual and ethical rigor. Their doctoral-level training brought a research-oriented perspective to data protection, patient consent, and staff privacy, promoting rigorous methodological approaches and comprehensive human subjects principles.

The technical workgroup, led by an EHR analyst, was responsible for system configuration, vendor integration, enabling the Ambient AI functionality within nursing documentation workflows, and security provisioning. Vendor team members supported configuration decisions and iterative refinements to align the technology with local nursing documentation practices. Vendor workgroup members also worked with operations team to implement the software as a smart-on-FHIR application into the EHR. While rigorous testing was completed by this team in staging environments, it is important to note that certain aspects of systems integration can only be evaluated within the live clinical environment, as not all technical integrations are fully replicated in testing or staging environments. Consequently, this approach may surface late–emerging findings that require timely operational response. In this project, implementation in the production environment revealed that unit–wide broadcast announcements intermittently interrupted recordings, a factor that could negatively influence user adoption. In response to this finding, nurse leaders and nursing staff discussed culture of non–essential broadcast practices, with the goal of minimizing recording interruptions and supporting more consistent use of the system.

The user experience/training workgroup, co–led by a nurse informaticist (CL) and a nursing director (KS), focused on workflow design, training materials, frontline usability considerations, and staff support strategies. Vendor representatives participated in the workgroup to advise on timelines, workflow constraints, and training resources. The training plan incorporated written communications and huddle points in the weeks preceding go-live, simulation training in a test environment, and nursing staff champion peer training. Consistent with nursing shared governance principles, the user experience/training workgroup regularly engaged the Unit Council to obtain feedback from frontline nursing staff. Discussions focused on workflow integration, perceived barriers, and opportunities for improvement. In addition, the health system provided protected time for a direct-care nurse (CG) to participate as a member of the research team. This role ensured integration of a frontline nursing perspective into training and implementation planning, including identification of anticipated staff questions and barriers, refinement of training content, and development of strategies to enhance staff receptivity to and adoption of the ambient AI tool.

The analytics workgroup, led by two nurse scientists, one embedded in the health system (JP) and one in academia (AW), was responsible for metric selection, dashboard development, and analytic approaches to evaluate documentation burden, adoption, professional well–being and system usability. Metrics were selected to reflect the multifaceted impact of the Ambient AI tool on inpatient nursing work and documentation burden, adoption, documentation quality, and user experience. Detailed metrics are provided in Figure 4. Consistent with an LHS approach, the evaluation emphasized objective, EHR–derived metrics complemented by validated survey instruments. Metrics were selected collaboratively by project stakeholders to ensure alignment with the operational mission, project charter, and clinical trial rigor. Ultimately, the clinical trial protocol served as a guide for operational deployment across data capture and primary and secondary metrics for clinical care. Dashboards were developed to support both ongoing operational monitoring and research evaluation. Data streams from operational systems and research data repositories will be transferred to the biostatistician for drift monitoring and analyses according to the statistical analysis plan. Additional analytic considerations include handling of staff who float between units and accounting for differences in adoption related to role as unit champions or training exposure.

**Figure 4.**
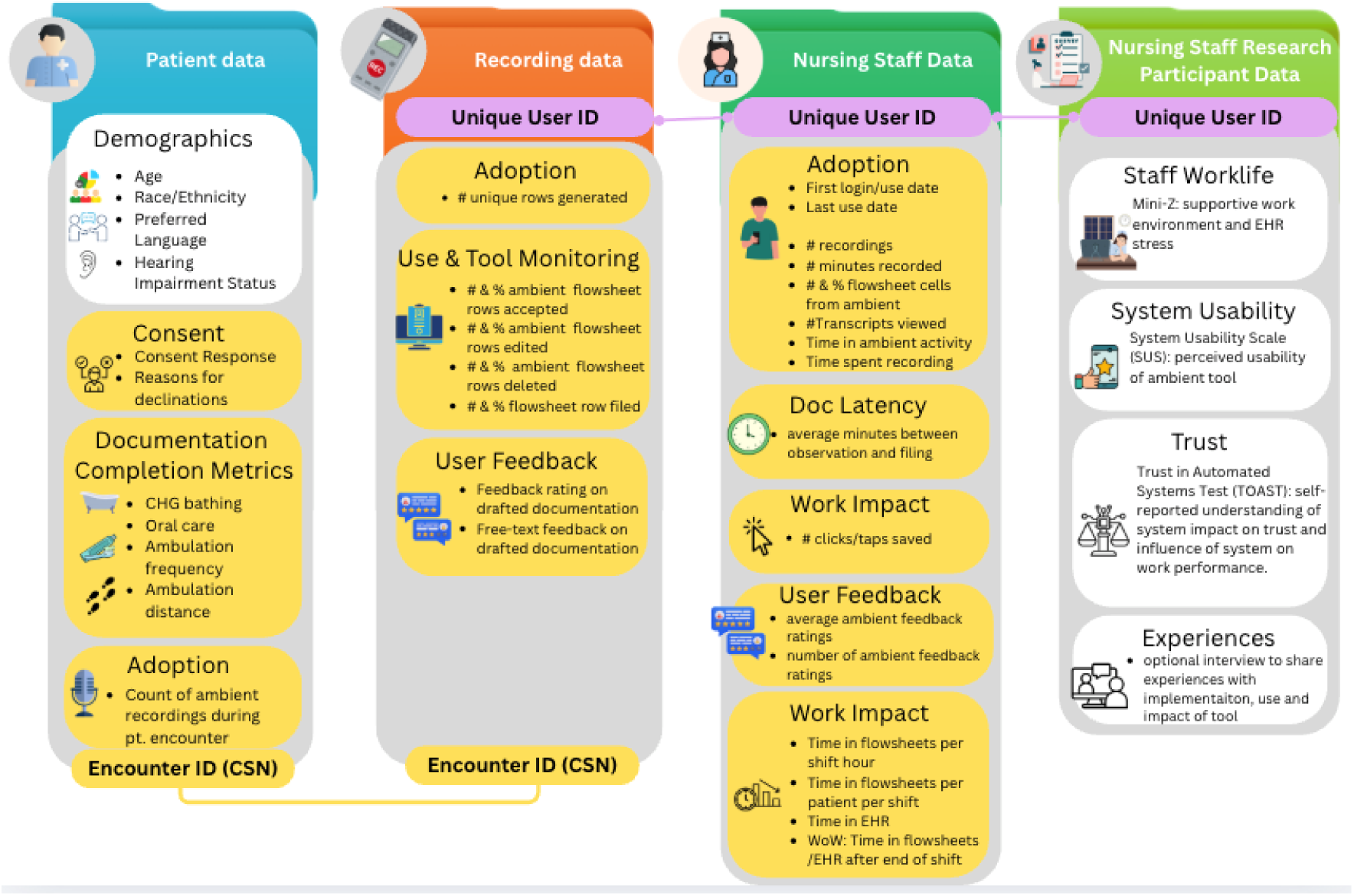
Infographic depicting metrics to be monitored and relationships between data tables. White background indicates metrics collected for research purposes.

The pragmatic trial was designed in November and December of 2025 and approved by the health system research operations committee in January of 2026. The IRB protocol was initially approved in April of 2026 with the most recent amendment approved in May 2026. The clinical trial was approved prior to the operational go-live data and informed the operational deployment.

## Discussion

Traditional efficacy trials are often poorly suited to evaluating clinical AI tools once deployed in practice, particularly those intended to operate continuously within complex sociotechnical environments [21]. Traditional controlled designs typically attempt to minimize noise through strict inclusion criteria and workflow standardization, which can obscure the very contextual factors that determine whether an AI intervention is effective in real-world care settings [22]. In contrast, the stepped–wedge pragmatic trial design used in this study was intentionally selected to accommodate real–world variability while still enabling causal inference, thereby reducing organizational uncertainty about whether observed changes could reasonably be attributed to the Ambient AI system rather than calendar time trends or unit–level confounding (Angus et al. 2025).

Balancing real–world flexibility with scientific rigor required explicit governance and communication structures, including close consultation with the study biostatistician to understand the analytic implications of changes required in response to operational needs. For example, the training strategy resulted in a group of nurse champions having access to the tool prior to broader unit–level exposure. This necessitated corresponding adjustments in the analysis plan to account for differential exposure and potential early–adopter effects. Another defining challenge of evaluating AI systems in real–world settings is that the technology itself may evolve during the project. In this trial, the vendor initiated a substantial model update during the planned rollout phase and recommended a delay in implementation to incorporate anticipated performance improvements, so the study calendar was adjusted accordingly. Making these explicit adaptations strengthens interpretability and offers practical guidance for other teams conducting AI evaluations under similar conditions.

While this trial was designed to inform local operational decision–making, it also contributes to broader reproducibility efforts in AI evaluation. By documenting both design decisions and implementation adaptations and posting key learnings and strategies to a public GitLab repository, this protocol provides a degree of standardization that can facilitate multisite comparisons without constraining necessary contextual tailoring. Such an approach supports cumulative knowledge generation across institutions, enabling health systems to learn not only whether Ambient AI technologies are effective, but under what conditions and for whom they are most likely to succeed.

Most existing Ambient AI implementation playbooks have been developed for outpatient provider (Physician and Advanced Practice Provider) workflows, which are characterized by bounded clinical encounters, primarily narrative documentation, and relatively controlled acoustic environments [23]. These assumptions do not translate well to inpatient nursing practice. Nursing work is distributed across multiple patients, unfolds continuously over shifts, and supports 24–hour care delivery rather than discrete visits. Documentation is predominantly structured, relying on discrete fields, dropdown selections, and “within defined limits” parameters that introduce different forms of cognitive and interactional complexity than narrative note creation. The inpatient nursing environment also presents substantially greater acoustic challenges for ambient technologies. Frequent alarms, overlapping conversations, visitors, televisions, and unit–level noise introduce conditions that are rarely encountered in outpatient exam rooms. These factors make nursing practice a particularly stringent test case for Ambient AI systems and necessitate evaluation approaches that explicitly account for environmental and workflow complexity.

Several elements of existing outpatient provider–focused Pragmatic Trial Operations (PTOps) playbook translated to nursing, including an LHS approach, core technical integration strategies, governance, and processes for iterative evaluation and alignment between operational and research goals [19]. However, substantial adaptation was required to address the realities of nursing work. For example, training and onboarding represented a significantly greater lift. Unlike many health practitioners who had prior experience working with human scribes, nurses were less likely to have experience verbalizing assessment findings. As a result, onboarding required more intensive support, including simulation–based training opportunities, to help nurses practice integrating conversational care into routine workflows. This represented not merely a technical transition, but a meaningful practice change in how documentation was produced during care delivery.

Governance structures addressing user experience, technical development, and analytics are foundational requirements for any Ambient AI implementation. However, existing outpatient provider Ambient AI playbooks often emphasize downstream documentation impacts related to coding and billing, reflecting health practitioner reimbursement models not applicable to nursing. By explicitly documenting which elements of existing playbooks transferred and which required reconfiguration, this protocol provides guidance for other institutions seeking to implement or evaluate Ambient AI technologies for nursing staff. Importantly, it highlights the risks of directly applying health practitioner –centric implementation models to nursing practice without sufficient adaptation.

The primary outcome of time spent in flowsheets per shift hour was chosen because it aligns with the operational goal of reducing documentation time. Use of an aggregated EHR–based metric also avoids reliance on nurse self–report, reducing nursing staff survey burden and enhancing scalability. However, this measure captures only one component of overall documentation workload and does not differentiate between documentation and data review. The latter may temporarily increase during early adoption as nurses need additional time to validate Ambient AI outputs. Time spent in flowsheets also does not assess documentation quality or changes in total documentation volume. As with many pragmatic AI trials, null or modest effects may reflect a learning curve with insufficient time for workflow stabilization rather than lack of impact. At the time of this publication, nursing staff champions on the first unit have begun using the Ambient AI technology in practice, providing essential data for model refinement and analytic dashboard validation. We currently have 16 participants enrolled in the survey data collection activities, demonstrating challenges of recruiting direct care nursing staff in study participation.

The protocol contributes to nursing-specific implementation and evaluation playbook, highlighting necessary adaptation to existing Ambient AI protocols that have largely focused on outpatient provider use. Through transparent reporting, open resources, and explicit documentation of governance and decisions, this trial aims to support both local operational decision making and broader reproducibility across LHSs. Aligning evaluation methods with operational decision–making needs enables practice-based evidence to support choices about whether to expand, modify, or retire AI technologies following initial piloting or deployment [22].

## Data Availability

All data produced in the present study are available upon reasonable request to the authors and all data produced are available online at https://git.doit.wisc.edu/smph-public/LearningHealthSystem/ambientlisteningnurseinpatient

## Acknowledgements

We would like to acknowledge Molly Gerhardt, Andrea McKenna, Jess Hunley, Anthony Stordalen, Alyssa Stauffacher, and many other members of the project team who were integral to the success of this project.

## Declaration of generative AI and AI-assisted technologies in the manuscript preparation process

During the preparation of this work the authors used Microsoft Copilot to assist with rephrasing some text segments for clarity and concision and to review alignment with journal formatting requirements. After using this tool, the authors reviewed and edited the content as needed and take full responsibility for the content of the published article.

